# BCG Vaccine Protection from Severe Coronavirus Disease 2019 (COVID19)

**DOI:** 10.1101/2020.05.05.20091975

**Authors:** Luis E. Escobar, Alvaro Molina-Cruz, Carolina Barillas-Mury

## Abstract

A series of epidemiological explorations has suggested a negative association between national BCG vaccination policy and the prevalence and mortality of COVID-19. However, these comparisons are difficult to validate due to broad differences between countries such as socioeconomic status, demographic structure, rural vs. urban settings, time of arrival of the pandemic, number of diagnostic tests and criteria for testing, and national control strategies to limit the spread of COVID-19. We review evidence for a potential biological basis of BCG cross-protection from severe COVID-19, and refine the epidemiological analysis to mitigate effects of potentially confounding factors (e.g., stage of the COVID-19 epidemic, development, rurality, population density, and age structure). A strong correlation between the BCG index, an estimation of the degree of universal BCG vaccination deployment in a country, and COVID-19 mortality in different socially similar European countries was observed (*r^2^* = 0.88; *p* = 8×10^-7^), indicating that every 10% increase in the BCG index was associated with a 10.4% reduction in COVID-19 mortality. Results fail to confirm the null hypothesis of no-association between BCG vaccination and COVID-19 mortality, and suggest that BCG could have a protective effect. Nevertheless, the analyses are restricted to coarse-scale signals and should be considered with caution. BCG vaccination clinical trials are required to corroborate the patterns detected here, and to establish causality between BCG vaccination and protection from severe COVID-19. Public health implications of a plausible BCG cross-protection from severe COVID-19 are discussed.

**Significance Statement:** The COVID-19 pandemic is one of the most devastating in recent history. The bacillus Calmette-Guérin (BCG) vaccine against tuberculosis also confers broad protection against other infectious diseases, and it has been proposed that it could reduce the severity of COVID-19. This epidemiological study assessed the global linkage between BCG vaccination and COVID-19 mortality. Signals of BCG vaccination effect on COVID-19 mortality are influenced by social, economic, and demographic differences between countries. After mitigating multiple confounding factors, several significant associations between BCG vaccination and reduced COVID-19 deaths were observed. This study highlights the need for mechanistic studies behind the effect of BCG vaccination on COVID-19, and for clinical evaluation of the effectiveness of BCG vaccination to protect from severe COVID-19.

## Introduction

The bacillus Calmette-Guérin (BCG) vaccine, an attenuated *Mycobacterium bovis* (1), has been extensively used in national vaccination programs, as it confers cross-protection from *Mycobacterium tuberculosis* infection (2). BCG vaccination of newborns and infants prevents disseminated childhood TB and reduces the risk of pulmonary TB by about 50% (3). Even though the BCG vaccine has been in use for more than 90 years, with proven safety, its efficacy is still controversial (4). BCG vaccination has shown clear protection in children, but in adults its effects have been inconsistent (5). Many countries initiated national BCG vaccination in the middle of the twentieth century with variable levels of coverage using different BCG strains (6), number of doses, and delivery method (7). As the prevalence of TB decreased, countries like France, Germany, and Spain stopped mass vaccination of children and moved to vaccinate only individuals at high risk. Other countries like Russia, Ukraine, and China have continued national BCG vaccination to date. Some countries never established national universal BCG vaccination, including the United States and Italy, and only target high-risk individuals (7).

There is ample epidemiological evidence that BCG vaccination has broad protective effects that are not specific to *M. tuberculosis* infection. For example, in 1927, Swedish children who received BCG vaccination at birth had a mortality rate almost threefold lower than unvaccinated children. This decrease of mortality could not be explained by TB infection, and thus early on, it was suggested that the very low mortality among BCG vaccinated children may be caused by nonspecific immunity (8). In West Africa, a BCG vaccination scar and a positive tuberculin reaction were associated with better survival during early childhood in an area with high mortality; this was not observed with other childhood vaccines (9). A general reduction in neonatal mortality linked to BCG vaccination was also reported in children from Guinea-Bissau, mainly due to fewer cases of neonatal sepsis, respiratory infection, and fever (10). The BCG vaccine appears to confer broad enhanced immunity to respiratory infections, as infants from Guinea-Bissau with acute viral infections of the lower respiratory tract were more likely to not have received BCG vaccination than matched controls (11). In Spain, hospitalizations due to respiratory infections in 0-14 year-old children not attributable to TB were significantly lower in BCG-vaccinated compared to non-BCG-vaccinated children (12). The observation that this protection was still present in 14 year-old children suggests that the broad protective effect of BCG can be long-lasting. Taken together, our current understanding of broad immune protection mediated by trained immunity and the epidemiologic evidence of long-lasting protection from viral infections of the respiratory tract, conferred by BCG vaccination, offers a rational biological basis for the potential protective effect of BCG vaccination from severe COVID-19 (reviewed recently (13)).

Considering the cross-protection reported for BCG vaccination on viral respiratory infections, recent publications have proposed that BCG vaccination could have protective effects against COVID-19 infection (14-16). These publications, however, do not include extensive statistical analysis, and the World Health Organization has cautioned about the lack of research regarding BCG vaccination against COVID-19 infection (17). In view of the growing interest to assess the plausible association between BCG vaccination and protection from severe COVID-19, we assessed available global data on BCG and COVID-19 to investigate the hypothesis that countries without a national BCG vaccination program would have greater COVID-19 mortality than countries that have a program. We attempted to control for potential confounding variables among countries such as, level of urbanization, population density, age classes, access to health, income, education, and stage and size of the COVID-19 epidemic.

## Methods

### Broad Analysis

We collected COVID-19 mortality data by country, considering that numbers of deaths may be more reliable than the numbers of cases, since case numbers are greatly influenced by the intensity of diagnostic efforts, criteria for testing, number of asymptomatic cases, and availability and sensitivity of diagnostic tests (18). Number of deaths are also a good proxy of epidemic size during explosive epidemics, considering that often the number of cases is underreported due to overwhelmed and unprepared health systems (i.e., *surveillance fatigue*) (19). Although media reports suggest consistent underreporting of COVID-19 deaths globally (20), this parameter is harder to manipulate than case numbers. We used ANOVA and t-tests to assess effect of BCG vaccination policy on COVID-19 mortality, and used linear regressions (21, 22) to assess linkages between use of BCG vaccine and number of COVID-19 deaths (*α*=0.05). BCG vaccination data were collected at the country level based on policy of vaccination (i.e., current, interrupted, never) (7, 23, 24) and the mean and median percentage of vaccination coverage during the period 1980-2018 (6), assigning zero coverage to countries where BCG national vaccination campaigns have never been conducted. COVID-19-related deaths were collected until April 23, 2020, and were standardized by population and stage of the epidemic (25). Independent (i.e., with vs. without BCG vaccination, policy of BCG vaccination in place, percentage of vaccine coverage) and dependent variables (i.e., mean, median, and maximum deaths per million) were compared using data at specific times of each country’s epidemic since the first death (e.g., 21 and 30 days since first death; days since first death until 0.1 and 1 death per million; middle-point and full period since first death). This design allowed fair comparisons between countries at different epidemic stage and of different population size, accounting for incubation period (26).

### Refined Analysis

The hypothesis that countries without a national BCG vaccination program would have greater COVID-19 mortality in adult populations than countries that have a program was investigated at the global level and filtered by social conditions. Analyses were performed initially for countries with BCG vaccination data, ≥1 M inhabitants, and nonzero COVID-19-related deaths. We focused on social variables as potential confounding factors, excluding climatic conditions in view of the lack of evidence for temperature-dependence of the COVID-19 epidemic (27, 28). Potential confounding variables were assessed based on access to health and education services and income (i.e., Human Development Index) (29), population size (30), human density (31), urbanization (32), and age structure of the population (33). For a fair comparison between countries, and after a preliminary evaluation of association (SI Appendix, Table S1), BCG vaccination and COVID-19 deaths assessments were conducted for countries with at least one death per million of inhabitants, ≥15% population with an age of 65 years or more, >60% of population living in urban areas, <300 inhabitants per km^2^, and a HDI >0.7. These inclusion criteria allowed comparisons of countries with similar social conditions to mitigated effects of confounding factors (SI Appendix, Table S1). To explore variables’ predictability, we developed multiple linear regression models between COVID-19 deaths accumulated during the first month of mortalities by country (log) and the potential confounding variables and BCG data. We used adjusted Bayesian information criterion, adjusted r^2^, and Mallow’s *Cp* metrics to determine the best variable combination and select the optimal model for COVID-19 death estimation. Analysis included all predictor variables combinations and were performed in R software using the *leaps* package (34). Considering the size and social disparities of the United States, BCG effect analyses were also performed considering the United States as a country, and separately by state.

At the regional level, we assessed the COVID-19 pandemic in the Americas assuming virus invasion by air traffic, as the pandemic presumably arrived in the American continent from Europe or Asia through this route. Air traffic has revealed a strong linear correlation between international COVID-19 cases and international passenger volume (*r*^2^=0.98, *p*<0.01) (35). In large countries, such as the USA, Mexico and Brazil, an epidemiological analysis at a country level does not consider the intense clustering of cases in states with large metropolitan areas and international airports, that were focal points of the pandemic. Thus, we decided to compare COVID-19 mortality in US states without BCG vaccination that have a high number of confirmed cases (more than 20,000 by 4/20/2020) with those states that were the main points of entry in Mexico and Brazil, as these two countries have current BCG vaccination programs. Evaluations were restricted to mortality 25 days after the first COVID-19 related death was registered. At the local level, we assessed the COVID-19 pandemic in Germany considering that East and West Germany followed different BCG vaccination schemes before the reunification in 1990. In West Germany, infants were vaccinated between 1961-1998, so that those 22-59 years old today were vaccinated (7). In East Germany, infants and 15 year-old teenagers with a negative skin test were vaccinated from 1951-1975; as a result, those 45-84 years old today received at least one dose of BCG (7). German states were compared accounting by age structure (36) considering that most of the mortality in Germany (95%) was observed in older individuals (60 years old or more) (37). We explored whether the broader range in BCG vaccination in eastern states of older individuals could reduce the mortality from COVID-19.

### Results

#### Coarse analysis

Global analysis of COVID-19 mortality revealed heterogeneity of mortality among countries (Figs. 1A). Three variables, HDI, percentage of population >65 yrs age, and urbanization, were consistently and positive associated with COVID-19 mortality (SI Appendix, Fig. S1), with the strongest association between human development index (HDI) and COVID-19 mortality (Figs. 1B; SI Appendix, Fig. S1). The global trend of vaccination policy (Figs. 1A; SI Appendix, Fig. S2) suggested an inverse association between BCG vaccination policy and COVID-19 mortality (Fig. 1C; SI Appendix, Fig. S3). Coarse analysis revealed a consistent link between BCG vaccination and COVID-19 mortality under different scenarios of data curation (SI Appendix, Tables S2-S4). For example, countries with a stronger BCG vaccination policy had significantly lower COVID-19 deaths/million (1 M) (SI Appendix, Table S2; Fig. 2A). More broadly, countries with current BCG vaccination had lower deaths as compared to countries with lack of, or interrupted BCG vaccination (SI Appendix, Table S3). Similarly, the percentage of BCG coverage was negatively associated with COVID-19 deaths/1 M (SI Appendix, Table S4). The association was consistently supported by the coarse-level analysis when considering the United States as a country or by analyzing individual states, and when correcting by the stage of the epidemic (SI Appendix, Tables S2-S4).

**Figure 1.**
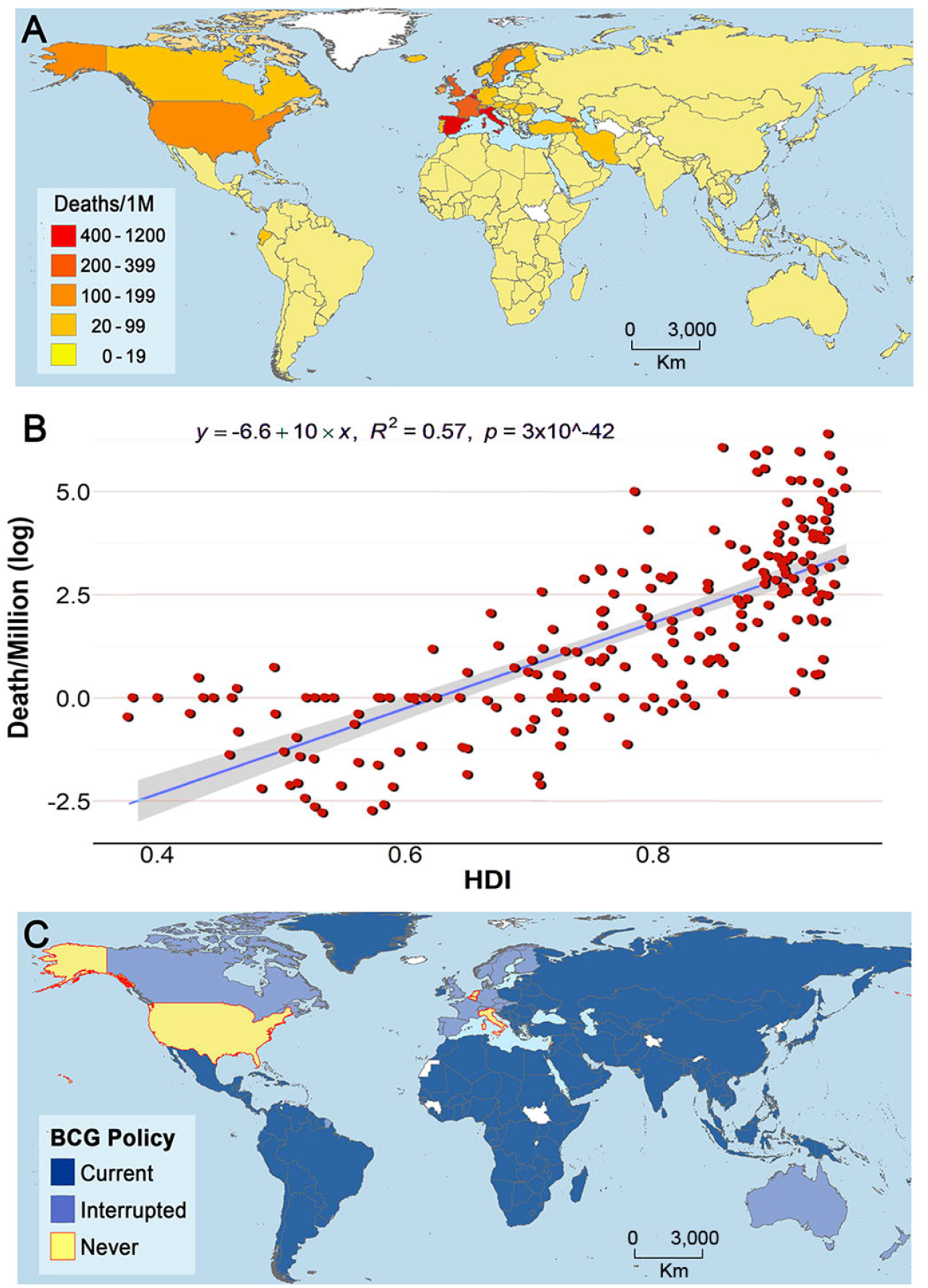
COVID-19 mortality, human development, and BCG vaccination policy by country. **A)** Map showing the COVID-19 mortality per million inhabitants in countries worldwide. D COVID-19-related deaths per country per 1,000,000 inhabitants denoting countries with low (yellow) to high (red) mortality. **B)** COVID-19 mortality per million inhabitants (log) vs. Human Developing Index (HID) in different countries worldwide. **C)** Map showing the BCG vaccination policy in countries that currently have universal BCG vaccination program (Current), countries with interrupted BCG vaccination programs (Interrupted), and countries that never implemented a universal vaccination program (Never). Countries without information appear in white.

**Figure 2.**
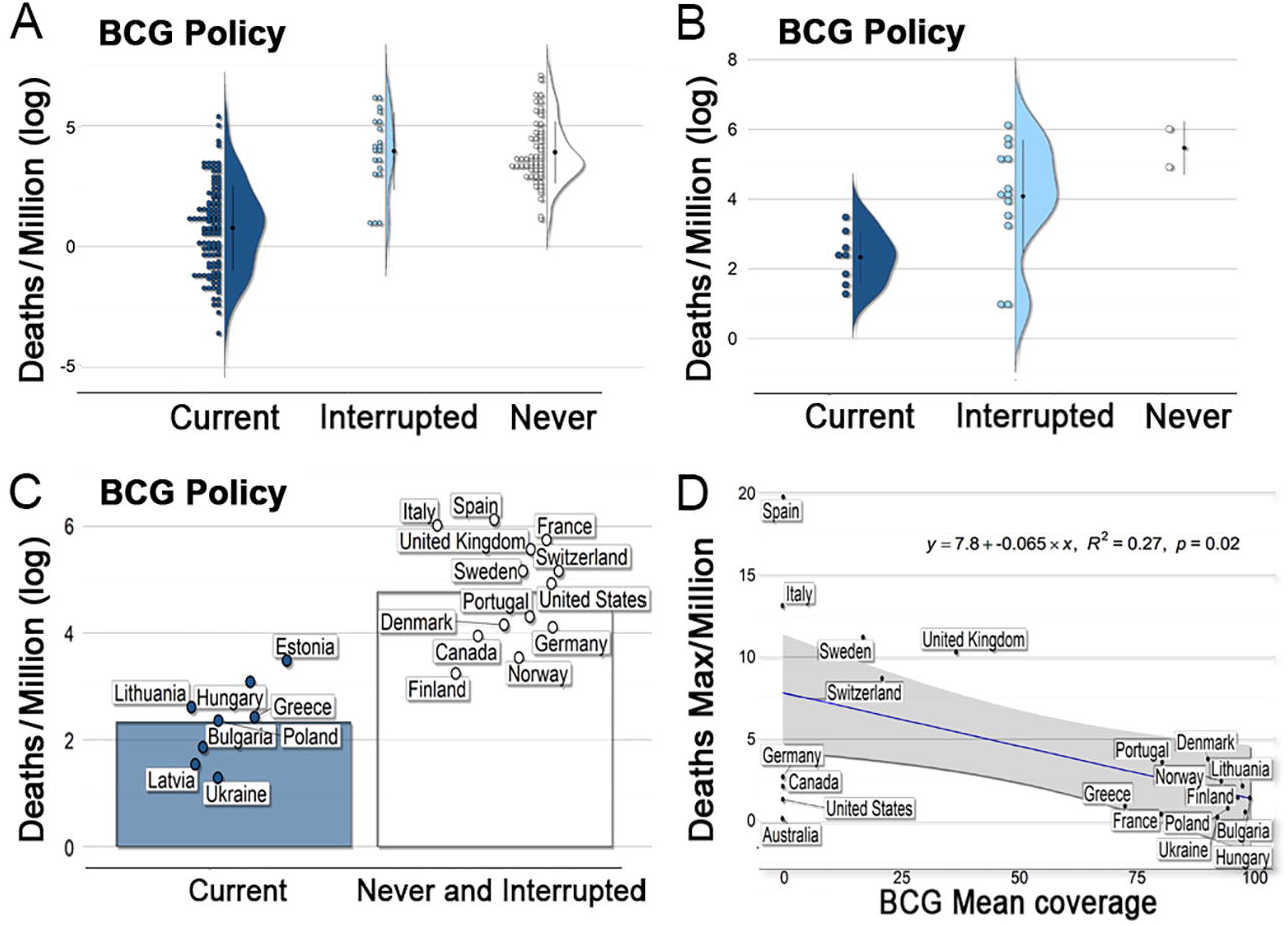
Linkage between COVID-19 mortality and BCG vaccination. **A)** Coarse analysis of COVID-19 mortality per million inhabitants in countries with current, interrupted or that never had BCG vaccination programs. **B)** Filtered analysis of COVID-19 mortality per million inhabitants in countries with current, interrupted or that never had BCG vaccination programs and similar social, economic, and epidemic stage conditions. **C)** Filtered analysis of COVID-19 mortality per million inhabitants in countries with current vs. interrupted or that never had BCG vaccination programs, including only countries with similar social, economic, and epidemic stage conditions. **D)** Negative association between percentage of vaccination coverage (mean) between 1980 and 2018, as a proxy of population protection, and maximum COVID 19 deaths per million inhabitants registered by country in a day, as a proxy of COVID-19 severity. Full list of analysis available in Supplementary Tables S2-S4.

#### Filtered analysis

When the effects of potentially confounding factors on the dependent variables were mitigated by including only countries with at least one death per million of inhabitants, ≥15% population with an age of 65 years or more, >60% of population living in urban areas, <300 inhabitants per km^2^, and a HDI >0.7 (SI Appendix, Table S1), we were left with 22 socially similar countries. The control of confounding variables reduced the overall significant effect and association between BCG and COVID-19 deaths (i.e., amount of significant evaluations for linear models went from 46% for coarse analyses vs. 19% for refined; ANOVA: coarse=100% vs. refined=31%; *t*-test: coarse=92% vs. refined=37%; Tables S2-S4). Nevertheless, a significant association and effect was still detected for several comparisons controlling for social conditions and stage of the epidemic by country (SI Appendix, Tables S2-S4). For example, type of BCG policy (i.e., current, interrupted, and never) had a significant effect on COVID-19 deaths/1 M (Fig. 2B; SI Appendix, Table S2), with a stronger policy being associated with lower COVD-19 mortality. Additionally, countries with current vaccination had lower deaths as compared with countries with lack of or interrupted BCG vaccination (Fig. 2C; SI Appendix, Table S3). Similarly, mean BCG coverage (%) was negatively associated with COVID-19 deaths/1 M reported by country during the first month of the pandemic (Fig. 2D; SI Appendix, Table S4). Our multivariate model revealed that HDI and population density had the best predictive capacity of COVID-19 deaths (*r*^2^=0.49, *F*(2,113)=55.57, *p*<0.001) (SI Appendix, Figs. S4A-C). It also showed that some of the variables evaluated were correlated with each other. For example, the percentage of population >65 yrs age and HDI are strongly correlated, while there is a moderate correlation between the time period of universal BCG vaccination and population density (SI Appendix, Fig. S4D).

#### Arrival of the Pandemic to the Americas

The COVID-19 pandemic arrived in the Americas by air traffic, and regions with high number of international flights from Europe have been the most affected. Thus, we decided to compare COVID-19 mortality in states from the United States without BCG vaccination with those that were the main points of entry in Mexico and Brazil, as these two countries have current BCG vaccination programs. We found that COVID-19 mortality in the states of New York, Illinois, Louisiana, Alabama, and Florida (unvaccinated) was significantly higher (*t*(237)=14.274, *p*<0.001) than states from BCG-vaccinated countries (Pernambuco, Rio de Janeiro, and Sao Paulo in Brazil; Mexico State and Mexico City in Mexico) (Fig. 3A, left panel). This is remarkable, considering that three states from Latin America have much higher population densities than the North American states analyzed, including New York (Fig. 3A, right panel).

**Figure 3.**
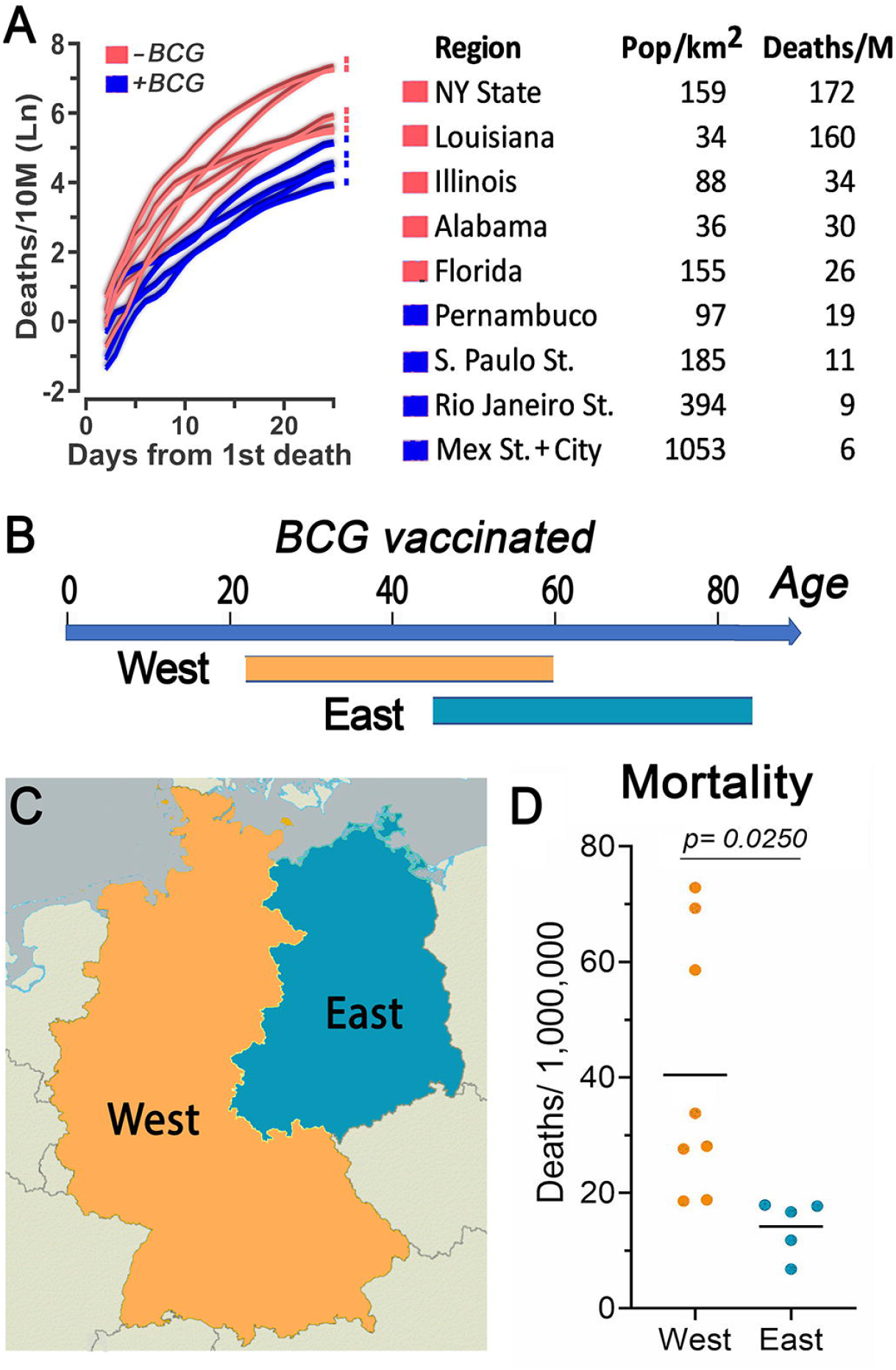
COVID-19 mortality in comparable regions that have had different BCG vaccination policies. **A)** COVID-19 mortality by time in populous North and South American States. COVID-19 mortality per 10 million inhabitants in a 3-day centered average. Time adjusted according to the day with the first death in each region as day 1, up to 25 days of the epidemic. Table shows population density and COVID-19 mortality by day 25 of the epidemic for each region. Regions that have had BCG vaccination (blue) had lower mortality than regions without BCG vaccination (red; *r*^2^=0.84, *p*<0.001; *t*(237)=14.274, *p*<0.001). **B)** Estimated age range of people that received BCG vaccination in East and West Germany. **C)** Map illustrating the regions of East and West Germany included in the analysis. D) Mean COVID-19 mortality in East Germany was lower than mortality in West Germany (*t*(11)=-2.592, *p*=0.025).

#### The Pandemic in Europe

We also found broad differences in COVID-19 mortality within Europe. Germany provides a unique opportunity to compare the potential effect of age of BCG vaccination on susceptibility to COVID-19, as, before the unification, East and West Germany followed different vaccination schemes. In West Germany those 22-59 years old today were vaccinated, while in East Germany those 45-84 years old today received at least one dose of BCG (Fig. 3B-C). A comparison of these two regions revealed that the average COVID-19 mortality rate in western German states (40.5 per million) was 2.9-fold higher than in eastern states (14.2 per million) (Fig. 3D). Similarly, the mean mortality in Western Europe was 9.92 times higher than in Eastern Europe (*t*(11)=-2.592, *p<*0.025) (Fig. 4A), where countries in general have active universal BCG vaccination programs (Fig. 4B).

**Figure 4.**
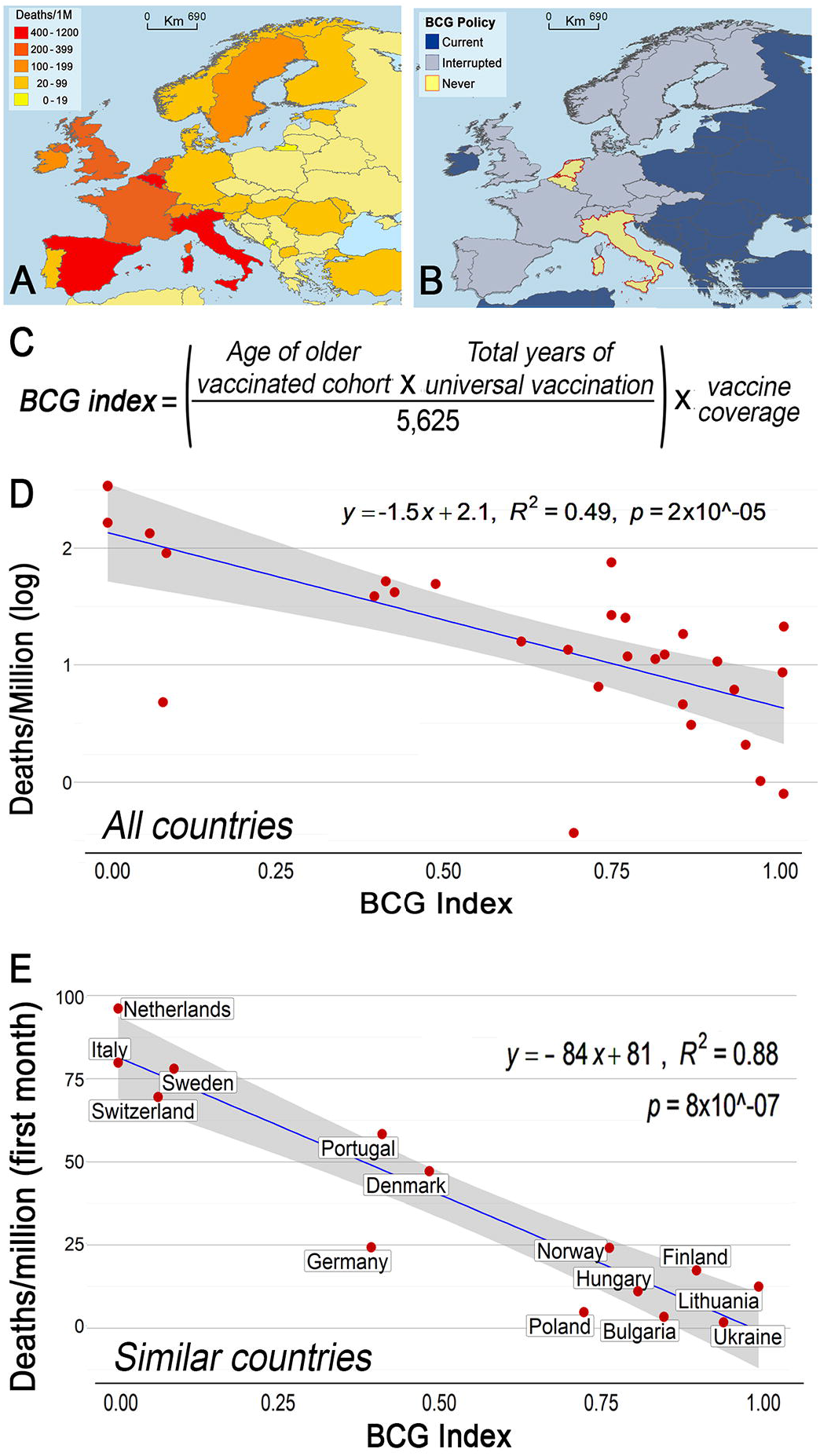
Variation in COVID-19 mortality and BCG vaccination policy in different European countries. **A)** Map illustrating COVID-19 deaths per million inhabitants in European countries. **B)** Map illustrating BCG vaccination policy in European countries that currently have universal BCG vaccination program (Current), countries that interrupted in the past a BCG vaccination program (Interrupted), and countries and never implemented a universal vaccination program (Never). **C)** BCG Index = (age of oldest vaccinated cohort x total number of years of vaccination campaign)/ standardization parameter (5625) X Mean BCG vaccination coverage. **D)** Correlation between BCG Index and COVID-19 mortality per million inhabitants and in European countries with different BCG vaccination policy. E) Correlation between BCG Index and COVID-19 mortality per million inhabitants during the first month of the pandemic in socially similar European countries with different BCG vaccination policy.

Western Europe has a complex history of BCG vaccination, ranging from Finland, which had a universal vaccination program for 65 years initiated in 1941, to other countries like Italy, Belgium and the Netherlands, which have never had universal BCG vaccination (Fig. 4B). To give these differences a quantitative value, a BCG vaccination index was developed by multiplying the age of the older group that was vaccinated (because COVID-19 mortality increases by age) by the number of years that a program was maintained. To adjust the index to a scale from 0 to 1, values were divided by a parameter of the maximum score expected (75 × 75 = 5,625, based on countries with 75 years of uninterrupted vaccination) Fig. 4C. We found a significant (*r*^2^=0.49, *p*<0.00001) negative correlation between BCG index and COVID-19 mortality in European countries that administer the BCG vaccine to infants by intradermal injection (Fig. 4D). In this analysis, the United Kingdom was excluded because it administered the vaccine to older children (12-13 years old) (7), and France was excluded because it administered the vaccine to infants and young children going to daycare, or to children of school age (38). We noted that COVID-19 mortality was higher in the United Kingdom and France compared to Germany, Scandinavia, or Eastern European countries (Fig. 4A). Finally, to try to control for other confounding factors that could affect COVID-19 mortality, we repeated the analysis including only those European countries that are socially similar (see *“filtered analysis”* section) and normalized the time of arrival of the pandemic by comparing the mortality during the first 30 days after the first COVID-19 death was reported in each country. A highly significant linear correlation was found between the BCG index and mortality during the first month of the pandemic (*r* = 0.88; *p* = 8×10^-7^) (Fig. 4E); indicating that every 10% increase in the BCG index is associated with a 10.4% reduction in COVID-19 mortality.

## Discussion

### Classic pathogen-specific immunity *vs* broad-protection through innate immune training

Cellular immunity is very important to control infections by intracellular pathogens, such as *M. tuberculosis*, responsible for tuberculosis disease (TB) in humans (39). Gamma interferon (*γ*-IFN) is a key cytokine produced by CD4+ T-cells that mediates macrophage activation and resistance to *M. tuberculosis* (40). Enhanced susceptibility to TB is seen in humans with mutations in the γ-IFN receptor (40) and in mice in which the γ-IFN gene has been disrupted (41). The BCG vaccine is thought to confer protection from TB by enhancing cellular immunity.

The innate immune system is the first line of defense against invading pathogens. Immune cells constantly patrol the different organs, especially the gastrointestinal tract, airways, and the lungs, which have direct contact with the environment. For many decades, the innate immune response was thought to be hardwired and unable to adapt and “learn” from previous exposure to a pathogen. In the last ten years several studies demonstrated that the priming or “training” of the innate immune system is an ancient response observed in evolutionary distant organisms including plants (42), insects (43), and humans (44). Trained immunity, defined as the enhancement in innate immune responses to subsequent infections, is achieved through epigenetic and metabolic programming of immune cells that allow them to mount a stronger response to pathogens and to activate adaptive responses more efficiently (45). Trained immunity can confer broad protection that is not pathogen-specific. For example, BCG vaccination is approved as a treatment for cancer of the bladder, and destruction of cancer cells has been shown to be mediated by trained immunity (46). Furthermore, BCG vaccination has been shown to elicit long-lasting innate immune responses, beyond those specific to mycobacteria (47), and to modify hematopoietic stem cells, resulting in epigenetically modified macrophages that provide significantly better protection against virulent *M. tuberculosis* infection than naïve macrophages (48). Taken together, our current understanding of broad immune protection mediated by trained immunity and the epidemiologic evidence of long-lasting protection from viral infections of the respiratory tract, conferred by BCG vaccination, offer a rational biological basis for the potential protective effect of BCG vaccination from severe COVID-19. This response to BCG vaccination appears to be mediated by a mechanism different from the cellular immune response that confer protection from systemic TB in children (13).

### BCG vaccination and COVID-19 mortality

The consistent association between BCG vaccination and reduced severity of COVID-19 observed in these and other epidemiological explorations is remarkable, but not sufficient to establish causality between BCG vaccination and protection from severe COVID-19. Randomized clinical trials, such as those ongoing in Holland (49) and Australia (50), in which health workers are administered either the BCG vaccine or a placebo saline injection, will determine the extent to which BCG vaccination in adults confers protection from COVID-19. There is limited information on the safety of administering BCG to senior persons, since BCG is a vaccine based on a live attenuated mycobacterium that should not be administered to immunocompromised individuals (51). *M. tuberculosis* infection can remain latent for decades and reactivate in the elderly when a senescent immune system loses the ability to contain the infection (52). Nevertheless, a small study found that vaccination of adults ≥ 65 years old with BCG prevented acute upper respiratory tract infections (53), and there is an active clinical trial vaccinating adults >65 years with BCG to boost immunity (54).

Most striking, COVID-19-related deaths are significantly higher in countries with higher quality of life (Fig. 1), contradicting the expectation of lower rates of mortality in countries with improved health care systems. Because of the differences in latitude, one can envision that differences in climate, such as ambient temperature, between states in the USA and South America could be responsible for the lower mortality in southern countries. However, one must consider that the average temperatures in the months of March-April are mild in Mexico City (62.2-64.6 °F), Sao Paulo (73.0-69.3 °F), Rio de Janeiro (77.1-73.9 °F), Florida (71.4-74.8 °F), Alabama (60.1-67.3 °F) and Louisiana (62.2-69.3 °F) (55), and that there is preliminary evidence for lack of temperature-dependence for the COVID-19 epidemic (27, 28). The augmented COVID-19 mortality in developed countries when compared to developing countries remained consistent even after correcting for social confounding factors and age. There are several important social differences between eastern and western German states, such as lower GDP in former East Germany, and higher percentage of population 65 or older in eastern states (24%) than in western ones (21%) (36), but it is hard to envision how these factors could decrease their risk of COVID-19 fatality in eastern German states. One should consider that the difference in mortality could also be explained, at least in part, by the lower population density in East Germany (154/Km^2^) than in West Germany (282/Km^2^). This variable is difficult to control, as there is great heterogeneity in population density even within each state.

The lack of apparent protection observed in the United Kingdom and France, where BCG vaccination was administered to older children, suggests that either trained immunity observed when infants are vaccinated is no longer achieved in older children, or it may be of shorter duration. There may be a “critical window” early in life where BCG vaccination can result in lifelong enhanced immune surveillance. It is also possible that the BCG strains used, or the administration route, also affect the innate immune response to vaccination. Considering the remarkable dissemination capacity and the mortality rates of COVID-19, vaccination capable of conferring even transient protection (e.g., 6-12 months) may be useful in individuals at high risk, such as health workers, first responders, and police officers, or those with pre-existing conditions such as obesity, diabetes, or cardiovascular disease. Similarly, even enhanced unspecific immunity through BCG vaccination in vulnerable age groups could ameliorate severe COVID-19. Temporarily induced trained immunity could buy time until specific vaccines and/or effective treatments against Severe Acute Respiratory Syndrome-coronavirus 2 (SARS-CoV-2) infections become available.

If the BCG protection hypothesis holds true, it would have great implications for regions with ongoing universal vaccination programs, including most developing countries, as they may experience lower morbidity and mortality during the pandemic than in Europe and North America. We found that the number of years that universal BCG vaccination has been implemented in a given country and the level of vaccinations coverage may play key roles in reducing of COVID-19 severity. For example, in many Latin American countries, universal vaccination was introduced in the mid 1960’s, suggesting that individuals ≥55 yrs would not be vaccinated and, in turn, represent a vulnerable segment of the population regarding COVID-19. Similarly, individuals born in years with low BCG vaccine coverage would be populations at risk. Most Asian countries have active universal BCG vaccination programs. If BCG is conferring some basal level of protection from COVID-19, it is possible that some of the social distancing roll-back strategies taken by Asian countries, in order to restart their economies, may not be effective in North America and Western European countries, and could result in a second wave of infections.

Our understanding of the biology of innate immune training is in its infancy (44, 45). Little is known about the capacity of BCG vaccination to confer broad immune enhancement and the functional correlates of protection. Our inability to confirm the null hypothesis of no effect of BCG on COVID-19 mortality could be explained by and alternative hypothesis of crossprotection mediated by BCG vaccination. We note, however, that the data used in this epidemiological study have important sampling biases, and that the statistical signal detected at the country level may not explain COVID-19 mortality at the local level. The possibility that a single exposure to an attenuated pathogen during infancy could result in life-long enhancement in immune surveillance is remarkable, but the available epidemiological data, in the absence of direct evidence from clinical trials, is not sufficient to recommend the use of BCG for the control and prevention of COVID-19 or other emerging infectious diseases.

## Data Availability

All data will become available upon publication

## Acknowledgments

The authors gratefully acknowledge Asher Kantor and Mark Johnson for editorial assistance and Mariana Castaneda-Guzman, Huijie Qiao, and Ana Beatriz Barletta Ferreira for assistance on data collection and analysis.

## Data Availability

All raw data were obtained from open sources and have been cited and deposited in the Supplementary Dataset file.

